# Medicaid Expansions and Private Insurance ‘Crowd-Out’ (1999-2019)

**DOI:** 10.1101/2023.05.22.23290306

**Authors:** Jason Semprini

## Abstract

Recent Medicaid expansions have rekindled the debate around private insurance “crowd-out”. Prior research is limited by short-time horizons and state-specific analyses. Our study overcomes these limitations by evaluating twenty years of Medicaid expansions across the entire United States. We obtain data from the U.S. Census Bureau for all U.S. states and D.C. for private insurance coverage rates of adults 18-64, for years 1999-2019. After estimating a naïve, staggered Two-Way Fixed Effects Difference-in-Differences regression model, we implement four novel econometric methods to diagnose and overcome threats of bias from staggered designs. We also test for pre-treatment differential trends and heterogenous effects over time. Our findings suggest that Medicaid expansion was associated with a 1.5%-point decline in private insurance rates (p < 0.001). We also observe significant heterogeneity over time, with estimates peaking four years after expansion. The importance of a 1-2% point crowd-out, we leave for future research and debate.

---

Since its inception in 1965 as a safety-net insurance program, Medicaid has steadily grown. With each decade, states have expanded the breadth of the program to cover more groups of children and adults^1–3^. Beginning as a program designed to fund healthcare services for people living with disabilities or in extreme poverty, the program now covers children, older adults, parents, and most recently working adults without children^4,5^. Since the 1990’s, nearly every state increased their Medicaid eligibility income thresholds. By expanding income eligibility over the years, working parents and childless adults gained access to low-to-no cost public health insurance^6,7^. Despite the obvious direct effect of increasing public health insurance coverage, the proliferation of Medicaid eligibility expansions have rekindled the “crowd-out” debate for private health insurance^8–11^.

The evidence regarding the impact of Medicaid expansion on private insurance is mixed and largely dependent on the specific context in which the expansion is implemented^12–19,19–21^. Some studies have suggested that Medicaid expansion has led to a considerable reduction in the number of individuals with private insurance^12–17^. Reducing the number of individuals who purchase private insurance could in turn lead to a reduction in revenue for private insurance companies, who then charge higher premiums, and ultimately causing a downward spiral of private coverage take-up^22,23^. There have also been reported impacts on quality of privately insured patient care and technology^24^. However, other studies have found that Medicaid expansion has had little to no impact on private insurance coverage^18–20^. Moreover, some studies have suggested that Medicaid expansion could have positive spillover effects into other health insurance markets^25,26^.

While the available evidence suggests a mixed impact of Medicaid expansion on private insurance, the short-term context and state-specific focus limit our ability to draw definitive conclusions. Further research with rigorous study designs and longer-term follow-up are necessary to fully understand the relationship between Medicaid expansion and private insurance crowd-out.

## Methods

### Data

The data used in this study were obtained from the Current Population Survey Annual Social and Economic Supplement^27^. For the purposes of this study, we used state-level data from the for the years 1999-2019. Specifically, we focused on the rates of individual, between ages 18-64, with private health insurance, as a percentage of the population aged 18 to 64 years.

Using a variety of publicly available sources, we then obtained information on state-level policies to identify when states expanded their income thresholds for Medicaid eligibility^6,28,29^. States that expanded after June 30 of the current year were considered treated in the subsequent year. Table 1 reports the expansion year for all states. Figure 1 reports the year-by-year private uninsurance rate trends by expansion status and year of expansion.

**Table 1:** State Medicaid Expansion Status. Table 1 reports the year each state expanded Medicaid income eligibility.

| **Expansion Year** | **States** |
| --- | --- |
| never | AL, FL, GA, ID, KS, ME, MS, MO, NE, NC, OK, SC, SD, TN, TX, UT, WI, WY |
| <1999 | DE, NY, VT |
| 2006 | MA |
| 2011 | DC, WA |
| 2012 | CA |
| 2014 | AZ, AR, CO, CT, HI, IL, IA, KY, MD, MI, MN, NV, NJ, NM, ND, OH, OR, RI, WV |
| 2015 | IN, NH, PA |
| 2016 | AK, MT |
| 2017 | LA |
| 2019 | VA |

**Figure 1:**
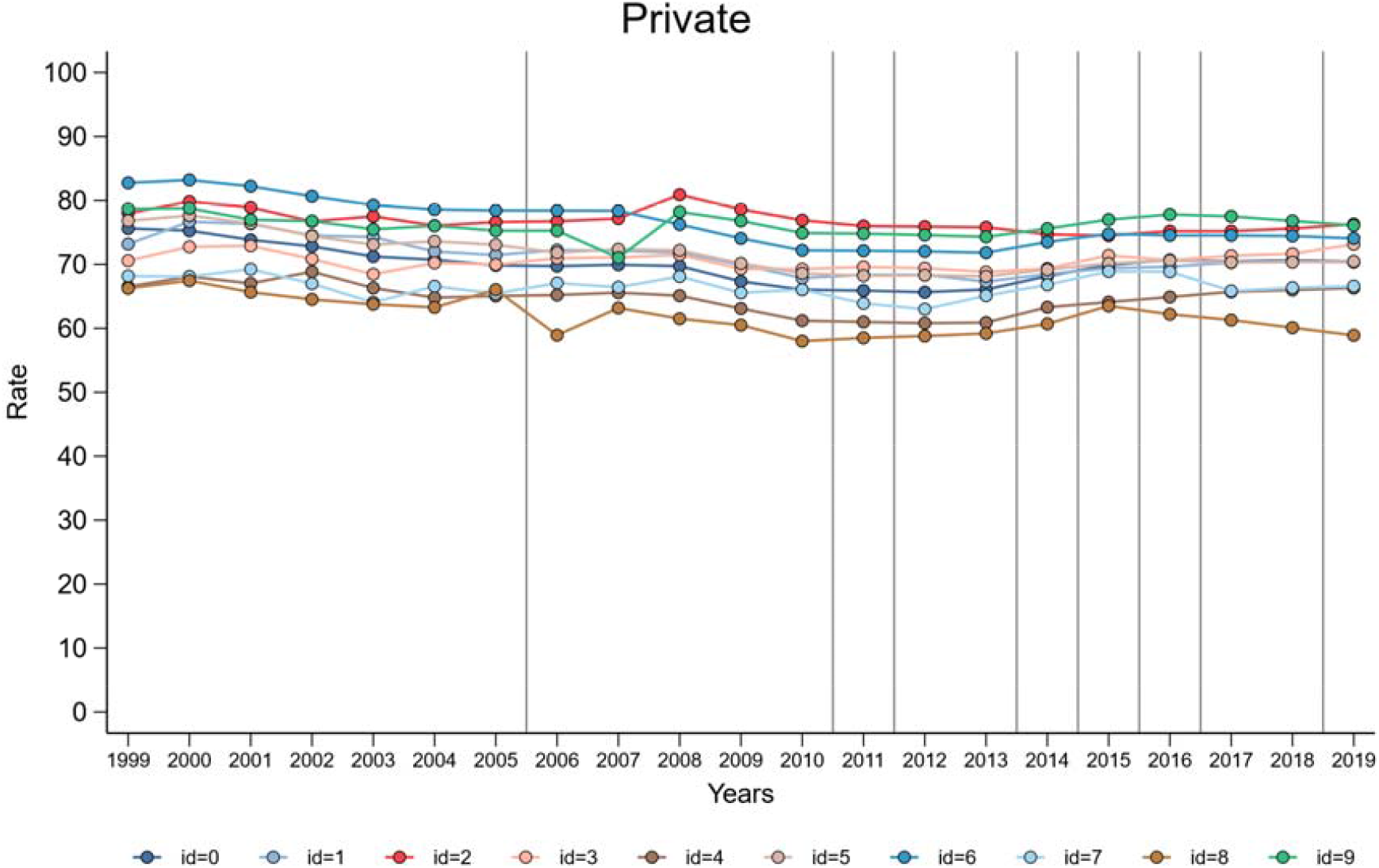
Private Insurance Rate Trends (1999-2019: Age 18-64) F1 visualizes the trends in private insurance rates (%) from 1999-2019 for adults between ages 18-64. Vertical grey lines indicate when each state (group ID) expanded Medicaid income eligibility. 0 = never (AL, FL, GA, ID, KS, ME, MS, MO, NE, NC, OK, SC, SD, TN, TX, UT, WI, WY); 1 = <1999 (DE, NY, VT) ;2 = 2006 (MA) ;3 = 2011 (DC, WA) ;4 = 2012 (CA) ;5 = 2014 (AZ, AR, CO, CT, HI, IL, IA, KY, MD, MI, MN, NV, NJ, NM, ND, OH, OR, RI, WV) ;6 = 2015 (IN, NH, PA) ;7 = 2016 (AK, MT) ;8= 2017 (LA) ;9 = 2019 (VA).

### Design

This study employs a Two-Way Fixed Effects Difference-in-Differences (TWFE-DD) design to examine the effects of state-level expansion of Medicaid eligibility on health insurance rates, specifically private insurance coverage^30^. We specify our reduced form equation here:

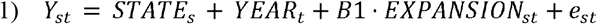

Equation 1 models Y = private insurance rate, for each state = s in year = t. By including state and year fixed effects, we compare state-level insurance trends for states that expanded Medicaid eligibility with trends in states that did not or have not yet expanded Medicaid. The primary null hypothesis is that expanding Medicaid in year >= t by state = s, was not associated with private insurance rates (B1 = 0). Our B1 estimate is unbiased if E(est) = 0, which would require an assumption that in the absence of expansion insurance rate trends would have remained parallel (common-trends) between states expanding and not expanding Medicaid^31^. This common trend assumption is satisfied if:

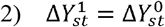

Equation 2 simply states that, in the absence of Medicaid expansion, the insurance rate trends in expansion states would be like the trends before and after the expansion years. The pre-treatment common trends assumption can be visually examined and empirically tested, but post-treatment common trends assumptions are unobservable counterfactuals.

#### Addressing Staggered Treatment

States did not randomly decide to expand Medicaid and did not all do so at the same time. Instead, state policy decisions related to Medicaid expansion were contextual and rolled out across states over time^32–34^. While a TWFE-DD design may be well-suited to address the potential endogeneity concerns associated with this type of policy exposure, recently scholars have exposed the valid concerns about the potential validity of staggered TWFE-DD designs in quasi-experimental policy research^35,36^.

##### Decomposition

After estimating a naive staggered TWFE-DD model, we first decompose the estimates with the Bacon Decomposition theorem. The Bacon Decomposition theorem proves that the estimand from a staggered TWFE-DD design is simply a weighted average of all 2×2 combinations of Difference-in-Differences (DD) estimates^35^. The weights are statistically determined by the relative share of each group and the group-level variance or timing of the group’s exposure to treatment. By understanding the sources of identifying variation within this staggered design, investigators can assess threats to the common-trends identification assumption and implement modern techniques to overcome threats to internal validity.

In unstaggered designs, heterogenous treatment effects are averaged over the entirety of the post-treatment period and do not necessarily bias the comparison between treatment and control groups after treatment. However, in staggered treatment designs, heterogenous treatment effects could bias all estimates where previously treated groups act as controls for later treated groups^35^. The bias is especially apparent if the treatment effects vary over time. For the staggered TWFE design, the common trend assumption in equation 2 must hold for all 2×2 DD’s, including both comparisons where expansion states act as respective controls.

The decomposed effect estimates and associated weights yield an estimate for the variance weighted average treatment effect on the treated. Here, B1 from equation 1 is unbiased under two conditions: Zero-Weighted Common-Trends and Time-Invariant Treatment Effects. If either condition fails, B1 is biased under traditional (naïve) staggered TWFE-DD designs. Equation 3 shows the potential sources of bias^35^:

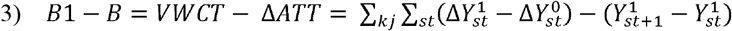

Equation 3 shows the two sources of bias. The first term on the right-side of equation 3 is the Variance Weighted Common Trends (VWCT). This term measures the sum of the counterfactual differential trends between each treated and control group estimates. Because this is unobservable, we cannot directly test if this term equals zero. However, we can use pre-treatment comparisons and the full set of weights to estimate the potential magnitude and direction of the VWCT bias term. First, note that the term is weighted (for all group = k and group = j comparisons). According to the Decomposition theorem, even if the unobserved common trend is non-zero, the bias goes to zero as the difference in weights approaches zero^35^. Regarding early and late expansion states, their respective common trend weight is the difference between their weight as a treatment group from their weight as a control group. The VWCT term can also equal zero in the presence of common trends, which can only be observed prior to treatment. The counterfactual common trend cannot be tested. Next, the dynamic or time-variant average treatment effects on the treated (ATT) is the sum of each group-specific change in ATTs over the course of the post-treatment period^37^. Even if the change in ATT does not vary by group, time-variant, increasing ATT biases (downward) the comparison between late-expansion states as treatment and early-expansion states as controls. Decreasing ATT biases the same comparison in the opposite direction.

##### Identification

In recent years, there has been growing interest in developing methods to overcome the limitations inherent with staggered TWFE-DD designs. In our study, we use three of the most popular methods for estimating causal effects with staggered TWFE-DD designs developed by Chaisemartin and D’Haultfoeuille, Callaway and Santanna, and 3) Borusyak.

The Chaisemartin estimator is a recent method that uses a “fuzzy” DD or Wald-DD principles, to estimate effects of a policy among groups changing their policy status (expanding Medicaid)^36,38^. This method overcomes any dynamic or time-variant effects and can also test for statistically significant non-common trends between expanding states and those states who have not yet expanded. Similarly, the Callaway and Santanna estimator overcomes potential bias from non-common trends and dynamic treatment effects by estimating effect parameters, not just by expansion status, but by group and cohort time periods^39^. The Callaway-Santanna method builds upon doubly-robust frameworks, which use inverse-propensity score weighting to match treatment and control groups, to estimate an effect of a policy across multiple dimensions and then aggregate the resulting estimates into a treatment effect measure. Finally, Borusyak’s method extends the theory and statistical technique of matrix imputation into the realm of unobserved potential outcomes^40^. Here, pre-expansion trends are used to impute “missing” or unobserved counterfactual insurance rate data to use as a comparison for estimating treatment effects and testing for non-common trends.

### Analysis

We analyze Equation 1 as a linear regression model with robust standard errors clustered at the state-level (except for the Callaway-Santanna method – where we instead estimated unclustered standard errors robust to heteroskedasticity). Bootstrapping procedures (999 reps) were used to account for time-variant dynamic treatment effects in the Chaisemartin method^36^. We visually examine our common-trends assumption using the Chaisemartin and Callaway-Santanna methods, and explicitly test for differential pre-treatment trends with all three methods as an event history analysis^36,39,40^. Statistical significance is set at alpha = 0.0125, using the Bonferroni method to adjust for multiple hypothesis testing^41^. All analyses were performed in STATA v. 17 using the xtreg, bacon_decomp, did_multiplegt, csdid, and did_imputation packages^42–46^.

## Results

The naïve TWFE-DD estimate would suggest that Medicaid expansion had no statistically significant association with uninsurance rates (Table 2). However, the decomposition suggests that the average result masks considerable heterogeneity by both ATT estimates and corresponding weights (Figure 2). Meanwhile, the results of the methods which account for bias inherent to staggered TWFE-DD designs suggest otherwise, with estimates nearly 50% higher in magnitude than the naïve estimate. The Chaisemartin method estimates that expansion was associated with a 1.517%-point decline in private insurance rates. The Callaway-Santana method estimates that expansion was associated with a 1.525%-point reduction. Finally, the Borusyak method estimates a 1.354%-point reduction. All three results were statistically significant (p < 0.001). These estimates correspond to a 2-3% relative decline in pre-expansion private insurance rates.

**Table 2:** TWFE-DD Estimates of Medicaid Expansion on Private Insurance Rates. Table 2 reports the results of the TWFE-DD linear regression models estimating the effect of Medicaid expansion on private insurance rates (1999-2019; adults between age 18-64). Standard errors are reported in parentheses. **** p < 0.001.

|  | Naïve | Chaisemartin | Callaway-Santana | Borusyak |
| --- | --- | --- | --- | --- |
| *ATT* | -0.985 | -1.517**** | -1.525**** | -1.354**** |
|  | (0.577) | (0.403) | (0.376) | (0.380) |

**Figure 2:**
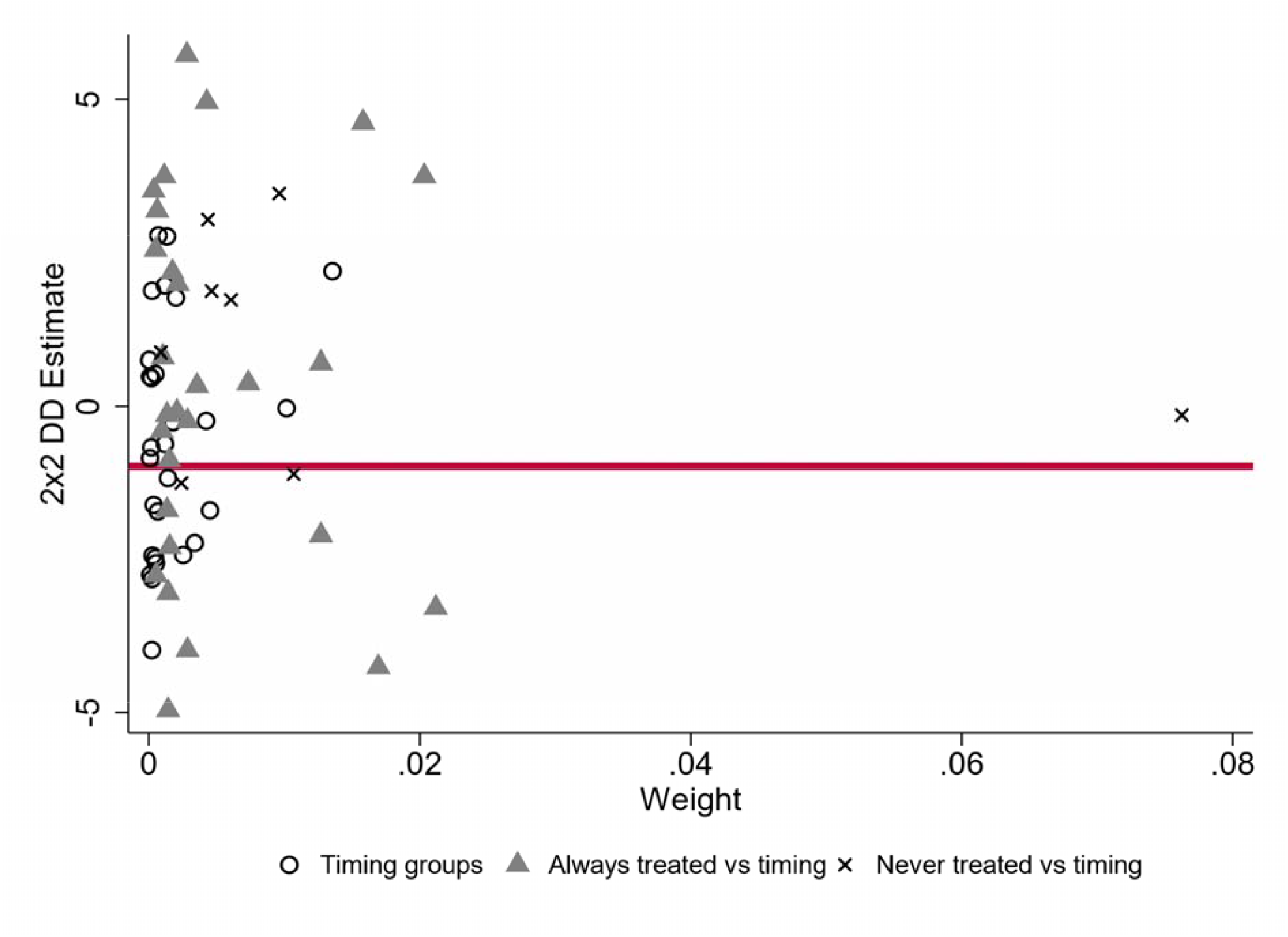
Decomposition of Naïve TWFE-DD Estimates of Medicaid Expansion on Private Insurance. Figure 2 reports the results of the Bacon Decomposition following the naïve TWFE-DD regression. The redline represents the overall DD Estimate = -.985 (p < 0.1). Timing group estimates involve previously expanding states as controls to expansion states.

We find no evidence that trends private insurance rates differed by expansion status in the years leading up to expansion. All pre-expansion coefficients are null and statistically insignificant (Table 3). These conclusion are consistent across all three of the modern TWFE-DD methods. We do, however, observe significant heterogeneity over time after expansion. Table 3 also reports the event-history estimates of expansion, for each year relative to the time of expansion. The estimates at year = T (year of expansion) range from 0.65% to 0.76%, and only statistically significant at p < 0.01 for two of the methods. At year = T+1 (year following expansion), estimates range from - 1.6% to -1.8% and are all statistically significant at p < 0.001. The estimates continue to rise, peaking at year T+4. Figures 3 and 4 depict the event-history heterogeneity. Finally, we do not find strong evidence that the reduction in private insurance rates differed by state. Except for states which expanded in 2006 (MA) and 2012 (CA), Figure 5 shows that Medicaid expansion was associated with significant reductions in private insurance for all states.

**Table 3:**
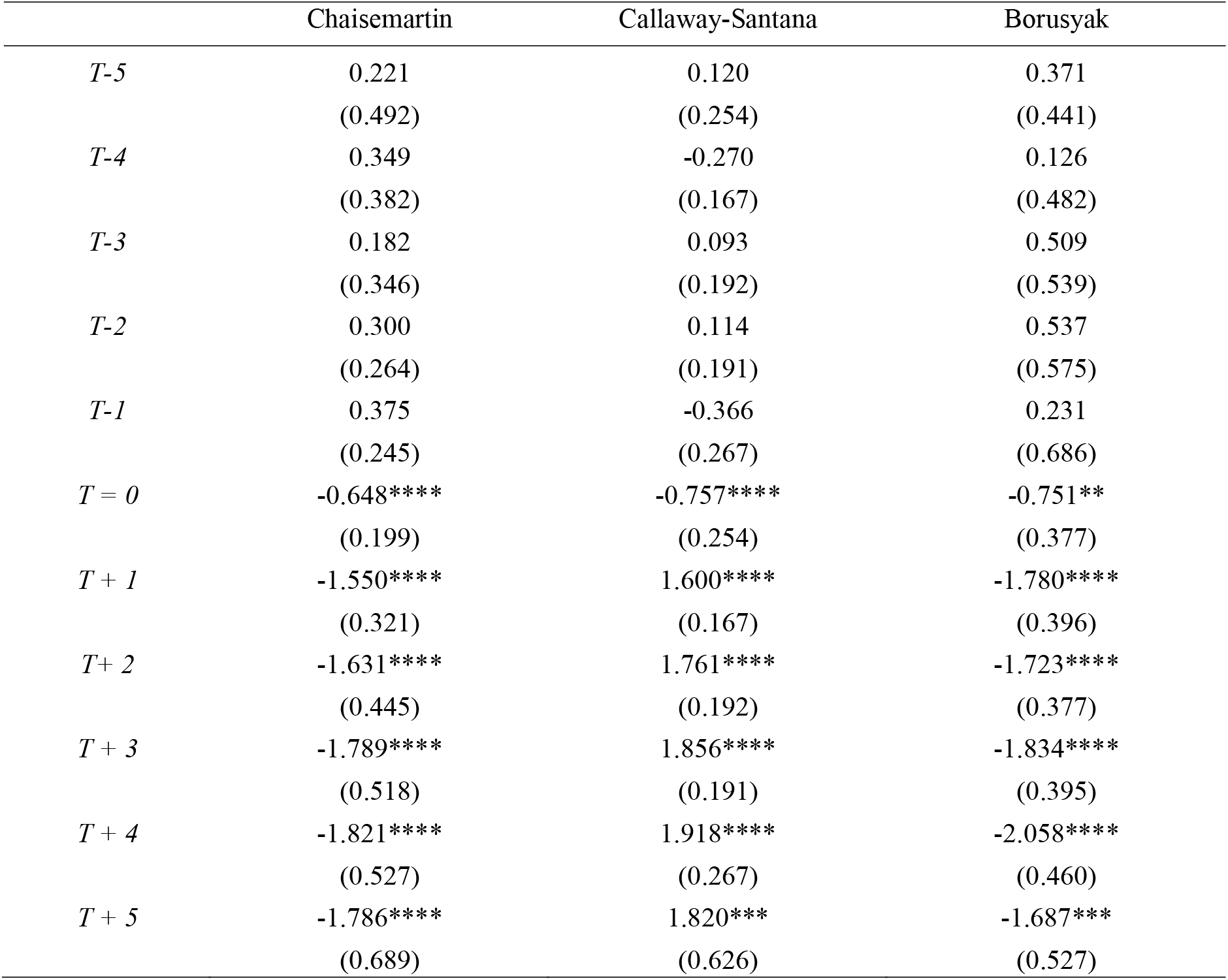
Event History Estimates of Relative Time to Medicaid Expansion on Private Insurance Rates. Table 3 reports the results of the Event History Analyses linear regression models estimating the pre-expansion (T-5 to T-1) differential trends and post-treatment (T = 0 to T+5) by expansion status on insurance (1999-2019; adults between age 18-64). Standard errors are reported in parentheses. T-5 indicates 5 relative years before expanding Medicaid; T-1 indicates 1 relative year before expanding Medicaid. T = 0 indicates year of Medicaid expansion. ** p < 0.05, *** p < 0.01, **** p < 0.001.

**Figure 3:**
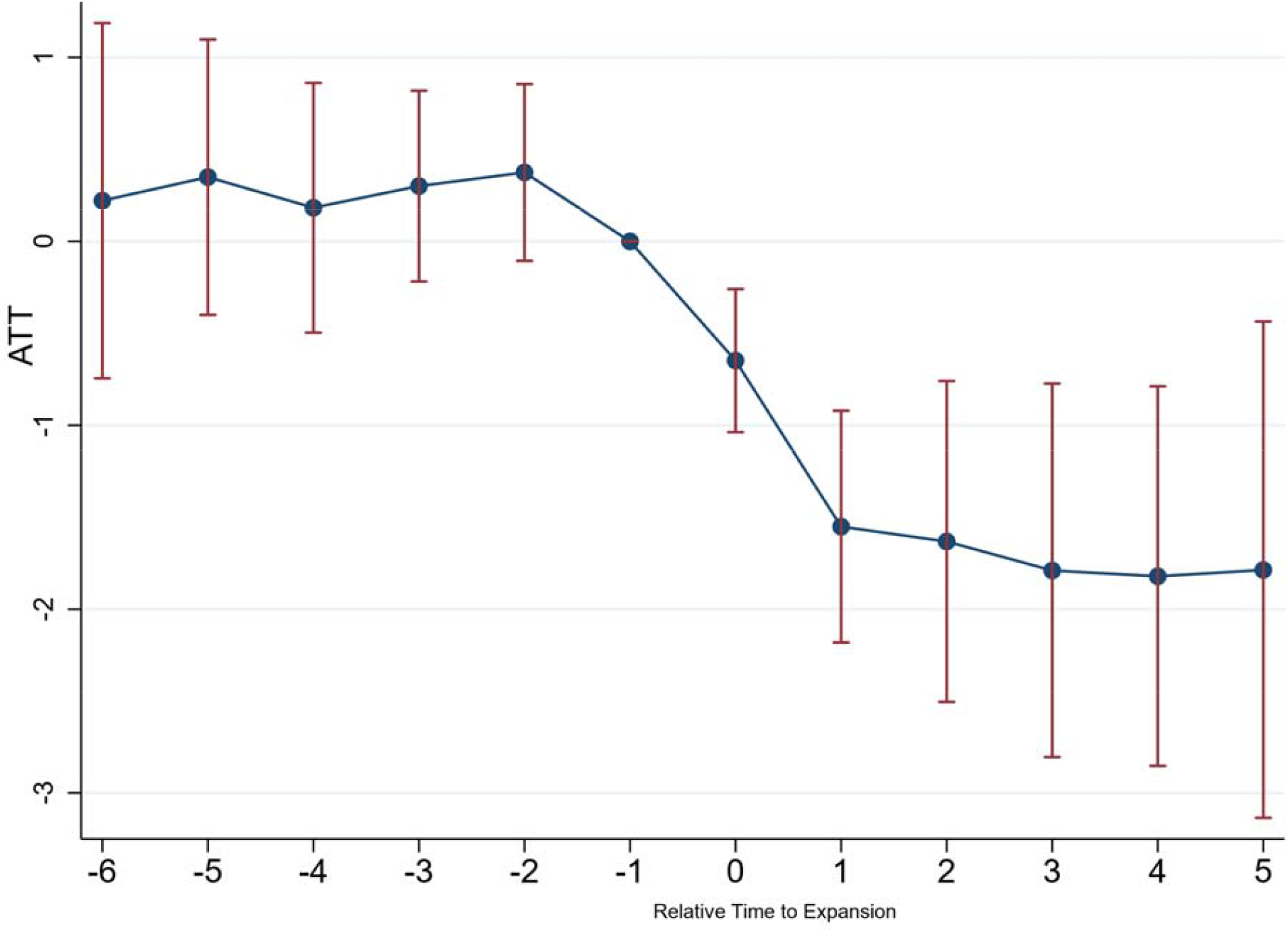
Event History Study Analysis of Medicaid Expansions Effect on Private Insurance. Figure 3 reports the Medicaid expansion coefficient estimates for each time-period relative to the year of expansion.

**Figure 4:**
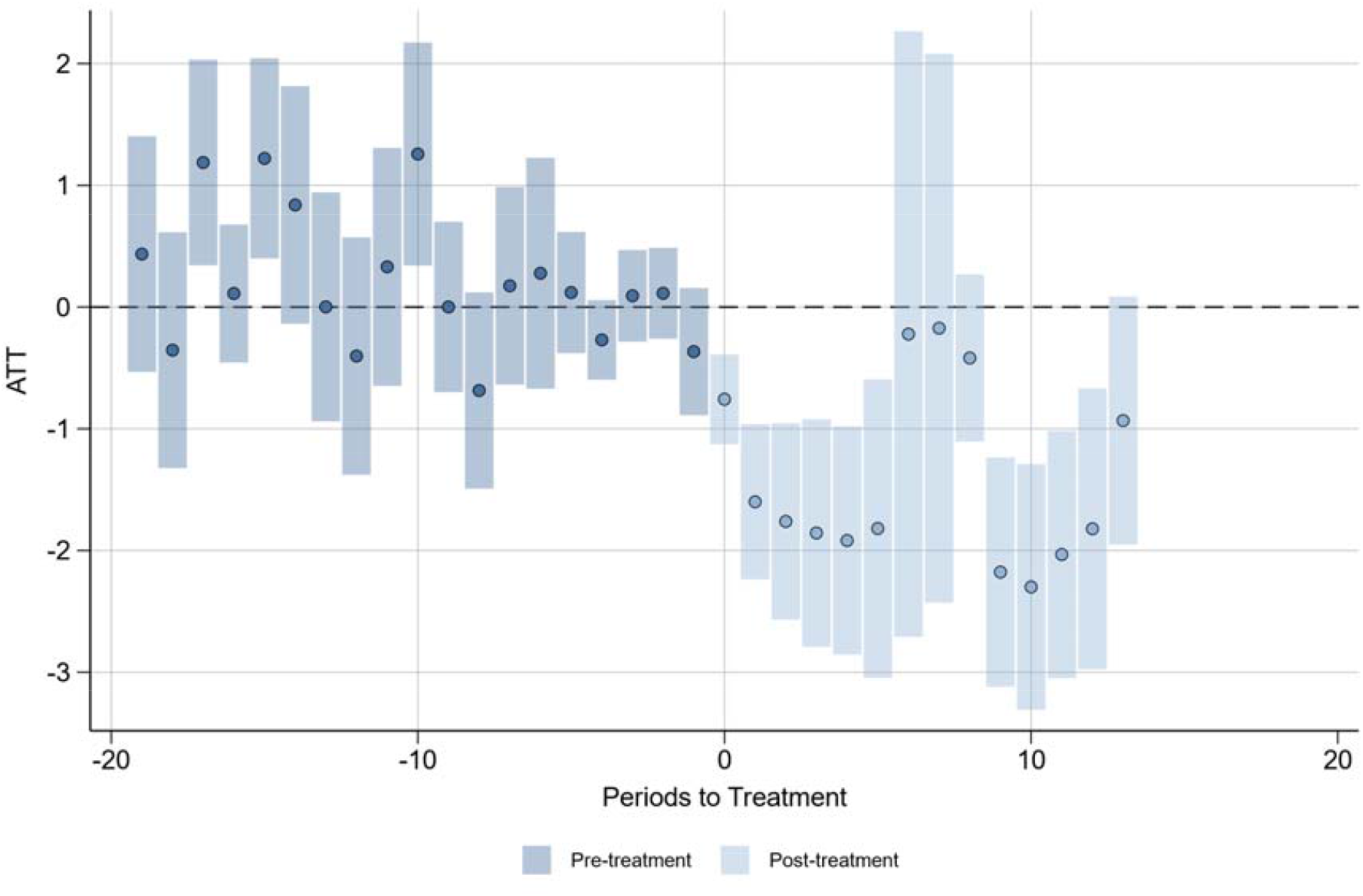
Event-History Analysis of Medicaid Expansions Effect on Private Insurance. Figure 4 aggregates the group and time-period estimates into an event-history analysis showing the differential trends by expansion status in private insurance before and after expansion.

**Figure 5:**
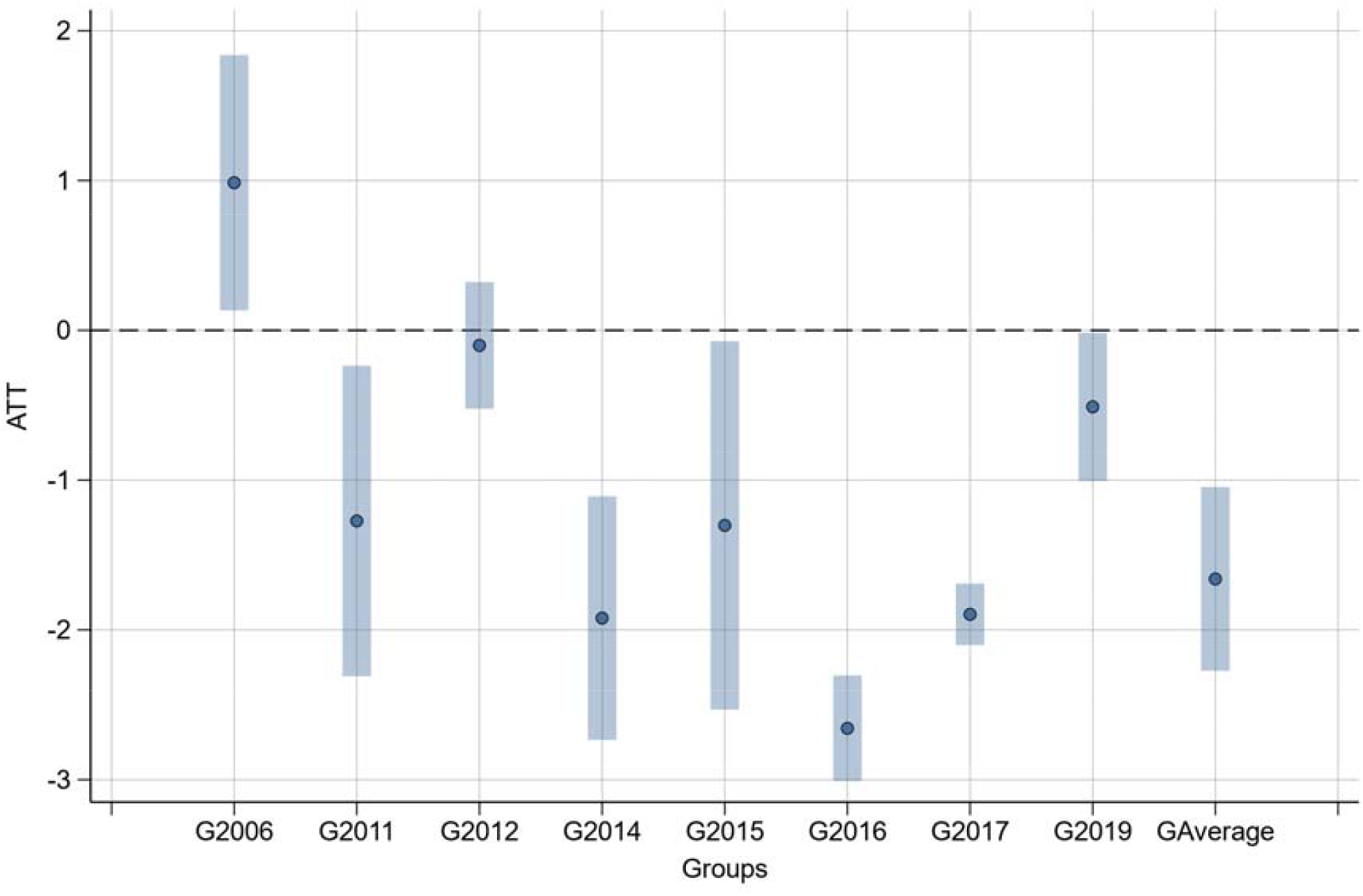
State Heterogeneity of Medicaid Expansions Effect on Private Insurance.

## Discussion

The findings of this study have significant implications for healthcare policy and practice, particularly regarding the expansion of Medicaid eligibility and its impact on private insurance rates. Our use of novel causal inference methodology allowed us to investigate a longer period and evaluate staggered expansions, accounting for time-variant dynamic and heterogeneous effects. Our approach allows us to overcome limitations present in previous studies and provide a more comprehensive understanding of the causal impact of Medicaid expansion on private insurance rates.

The key implication of our study is the observed crowding-out of private insurance rates caused by Medicaid expansion. While the decrease in private insurance rates may result in savings for individuals, an increase in Medicaid enrollment and associated costs may offset these savings. Therefore, careful analysis of the budgetary impact is necessary to assess the overall financial implications of expanding Medicaid eligibility^47^. With more individuals opting for Medicaid, the burden of providing healthcare services and associated costs will continue shifting from the private to the public sector^48^. The shift from private to public financing for insurance contrasts the increasingly privatized management of government insurance^49^. This redistribution can have both positive and negative implications, depending on the distributional impacts and overall efficiency of the healthcare system.

Moreover, our study investigates heterogenous effects of expansion. Understanding that the impact of Medicaid expansion may vary across different populations and contexts is essential for designing targeted policies and assessing the distributional consequences of expansion. By examining heterogeneity, we shed light on how expansion affects private insurance rates. We found that the crowd-out effect of Medicaid expansion peaked between 2-5 years after expansion, then leveled off or diminished. We did not, however, observe significant state heterogeneity. With the exception of Massachusetts, all other average state effects from Medicaid expansion reduced private insurance rates. Knowing that

## Limitations

Our study has several limitations. First, the use of state-level data restricts the granularity of analysis and may overlook within-state variations. Second, the absence of control variables, chosen to maximize flexibility and limit bias from arbitrary analytic decisions, limits our ability to account for other influencing factors. Third, being an observational study, we cannot establish a definitive causal relationship, despite employing advanced causal inference techniques. Endogeneity concerns persist, despite rigorous econometric techniques employed to mitigate them. Future research should address these limitations through more granular data and subgroup analyses to enhance the robustness and generalizability of our findings.

## Conclusions

Medicaid expansions reduced private insurance rates. Yet, we cannot ascertain how this crowd-out may have affected consumer welfare or government budgets. Recognizing the crowd-out, however, enables policymakers to make informed decisions regarding healthcare policy. Future research and policymaking should consider this trade-off. We leave discussion of the importance of a 1-2% point crowd-out for future work.

## Data Availability

All data and analytic code are available at author's publicly available GitHub repository: https://github.com/jsemprini/medicaidexpansion_crowdout

https://github.com/jsemprini/medicaidexpansion_crowdout/blob/main/stata-state-1999-2019.dta

